# A joint hierarchical model for the number of cases and deaths due to COVID-19 across the boroughs of Montreal

**DOI:** 10.1101/2021.10.06.21264645

**Authors:** Victoire Michal, Leo Vanciu, Alexandra M. Schmidt

## Abstract

Montreal is the epicentre of the COVID-19 pandemic in Canada with highest number of deaths. The cumulative numbers of cases and deaths in the 33 areas of Montreal are modelled through bivariate hierarchical Bayesian models using Poisson distributions. The Poisson means are decomposed in the log scale as the sums of fixed effects and latent effects. The areal median age, the educational level, and the number of beds in long-term care homes are included in the fixed effects. To explore the correlation between cases and deaths inside and across areas, three bivariate models are considered for the latent effects, namely an independent one, a conditional autoregressive model, and one that allows for both spatially structured and unstructured sources of variability. As the inclusion of spatial effects change some of the fixed effects, we extend the Spatial+ approach to a Bayesian areal set up to investigate the presence of spatial confounding.

## 1. Motivation

As of July 25th, 2021, the coronavirus disease 2019 (COVID-19) pandemic counts a total of 194,248,750 cases and 4,163,599 deaths worldwide (Dong et al., 2020). This disease is caused by an infection with the severe acute respiratory syndrome coronavirus 2 (SARS-CoV-2). The World Health Organization declared COVID-19 to be a pandemic in March 2020 (Ghebreyesus, 2020) and since then, countries worldwide have instituted infection control measures (e.g., lockdowns, curfews, mask mandates) to control the spread of the disease. We are interested in studying the COVID-19 spread in the urban agglomeration of Montreal. As of July 25th, 2021, Canada cumulates 1,426,903 cases and 26,547 deaths due to COVID-19 (Government of Canada, 2021) with 9.2% of the cases and 17.9% of the deaths recorded in Montreal ([dataset] Direction régionale de santé publique, 2021). The agglomeration covers approximately 500 km^2^ and is the most populous administrative region in the province of Quebec, Canada, with over 1.9 million inhabitants which represents, approximately, 6% of the Canadian population (Statistics Canada, 2017a,b). The agglomeration incorporates 19 boroughs of the City of Montreal and 15 related cities. These boroughs and related cities vary in demography, socio-economic status, and healthcare infrastructure.

This analysis focuses on data on the cumulative number of COVID-19 confirmed cases and deaths in Montreal since the beginning of the pandemic, as of July 25th, 2021. The cases and deaths distributions across the areas of Montreal are shown in Figure 1. This dataset is made publicly available by the regional director of public health ([dataset] Direction régionale de santé publique, 2021). The data consist of 34 boroughs and related cities of Montreal. However, one related city, L’Île-Dorval, did not record cumulative data. We exclude this area from the analysis, yielding a total of 33 areas.

**Figure 1:**
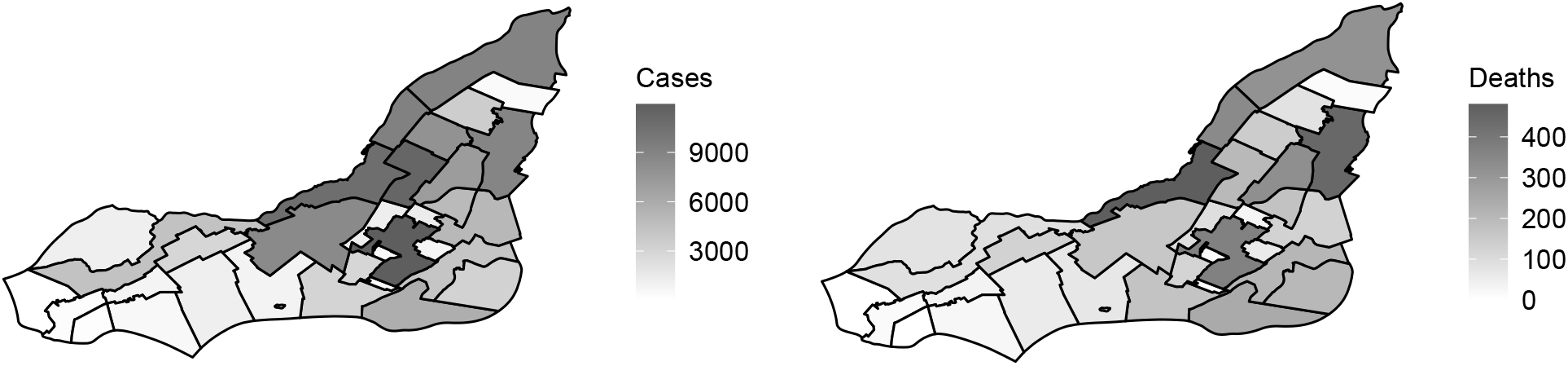
Maps of the COVID-19 cases (left) and deaths (right) across the 33 boroughs of Montreal.

The Ministry of Health and Social Services defines a confirmed case or death due to COVID-19 as follows. Cases are confirmed either by a laboratory or by an epidemiological link. A laboratory confirms a case through the detection of nucleic acids of SARS-CoV-2. To confirm a case by an epidemiological link, the manifestation of clinical symptoms must be compatible with COVID-19 with a high risk of exposure to a case that was confirmed by a laboratory during the period of contagion and no other apparent cause. Deaths may also be confirmed by a laboratory or by an epidemiological link. A laboratory confirms a death due to COVID-19 through the manifestation of clinical symptoms compatible with COVID-19 before death and the detection of nucleic acids of SARS-CoV-2. A death is confirmed to be due to COVID-19 by an epidemiological link if there was a manifestation of clinical symptoms compatible with COVID-19 before death with a high risk of exposure to a case that was confirmed by a laboratory during the period of contagion and no other apparent cause (Ministère de la Santé et des Services sociaux, 2021). The death counts of two areas are censored for privacy issues. Namely, Baie-d’Urfé and Montréal-Ouest recorded between 1 and 4 deaths. We also obtained data on the population size of each borough and related city from the 2016 Canadian Census ([dataset] Ville de Montréal, 2016).

In the literature, many studies identify age as a risk factor of COVID-19 mortality (Zhou et al., 2020; Bonanad et al., 2020; Yanez et al., 2020). Some studies also highlight the relationship between socio-economic status and COVID-19 cases (Hawkins et al., 2020; Gangemi et al., 2020) and COVID-19 deaths (Hawkins et al., 2020; Bermudi et al., 2021). Moreover, apart from COVID-19, there exist important health disparities due to socio-economic inequalities in Montreal. These have been shown to lead to a difference in life expectancy of up to 11 years between areas (Le Blanc et al., 2011). On the other hand, healthcare infrastructure has been positively associated with reactive responses to COVID-19 (Sharma et al., 2021). In Quebec, there exist five classes of senior residences. Three of these fall under the denomination of residential and long-term care centres, which may be public, private and subsidised by the government, or private and non-subsidised. Most senior residences are of the two remaining classes: intermediary resources and private seniors’ residences. Although all of these residences can provide similar services, residential and long-term care facilities specifically serve seniors with an important loss of autonomy (Gouvernement du Québec, 2021). Hospitals in Quebec, under the direction of the Ministry of Health, were generally better prepared to respond to the COVID-19 pandemic than long-term care homes during the first months of the pandemic (Doucet, 2020). The vast majority of COVID-19 deaths in senior residences in Quebec are linked to long-term care homes. In fact, as of July 25th, 2021, 51% of all COVID-19 deaths in Quebec are linked exclusively to residential and long-term care centres (Institut national de santé publique du Québec, 2021).

This literature review motivates our covariates’ selection in order to model the cases and deaths due to COVID-19 in the 33 areas of Montreal. To account for the age profile of each borough, we obtain data on the median age of the population by borough and related city from the 2016 Canadian census ([dataset] Ville de Montréal, 2016). As a proxy for the socio-economic status of each borough, we consider the percentage of the population between 25 and 64 years old with a university diploma. These data are also extracted from the 2016 Canadian census ([dataset] Ville de Montréal, 2016). Finally, regarding the healthcare infrastructure in Montreal, we include the number of beds in all long-term care homes of each borough (*Centre d’hébergement et de soins de longue durée*, CHSLD). These records are available in public databases ([dataset] Ministère de la Santé et des Services sociaux, 2021a,b).

In this paper, we analyse the *joint* distribution of COVID-19 cases and deaths across the 33 boroughs of Montreal through a Bayesian hierarchical joint model. We are interested in measuring the strength of the associations between the three available covariates and the cases and deaths due to COVID-19. Also, we aim at investigating how these two outcomes are correlated within and across boroughs. We examine whether the covariates impact the risk of being a case or the risk of dying from COVID-19 in a similar fashion. Additionally, we investigate if there is any structure left in the data after accounting for the available covariates. We explore different prior specifications for these local latent effects, ranging from independent to spatially structured ones. It is known that the inclusion of spatially structured latent effects can affect the estimation of the fixed effects (Reich et al., 2006; Khan and Calder, 2020; Dupont et al., 2021); this is known in the literature of Spatial Statistics as spatial confounding. We fit a Bayesian version of the *Spatial+* approach proposed by Dupont et al. (2021) to investigate if there is spatial confounding in the fitted models.

This paper is organised as follows: Section 2 describes the proposed models, the method for spatial confounding adjustment and the inference procedure. In Section 3, the results of the analyses of the COVID-19 cases and deaths are presented. Finally, Section 4 concludes and discusses our findings.

## 2. Methods

Let *C*_*i*_ be the recorded number of COVID-19 cases in borough *i*, for *i* = 1, …, *n*, where *n* = 33 is the number of non-overlapping boroughs in Montreal. Let *D*_*i*_ be the number of deaths due to COVID-19 in borough *i*. We model the deaths and cases through a joint hierarchical Bayesian model in order to accommodate the natural relationship one expects between cases and deaths of a disease. First, we define a marginal distribution for the cases, and then, conditional on the number of cases, we define a distribution for the number of recorded deaths. Sahu and Böhning (2021) modelled the cases and deaths due to COVID-19 in England in a similar fashion, using a 2-stage Bayesian hierarchical model. However, their interest lied in the temporal structure of the data and did not consider joint latent effects. Our approach differs because we do not conduct a spatio-temporal analysis, and we are interested in measuring the correlation between cases and deaths inside and across the boroughs of Montreal. To that end, we include a correlation parameter that helps borrow strength from the cases and deaths inside and across boroughs. More specifically, we assume,

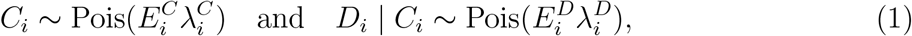

where Pois stands for the Poisson distribution, 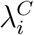 and 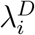 denote, respectively, the relative risk of the cases and the deaths in area *i*, whereas 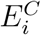 and 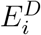 are offsets. The offset for the number of cases is computed based on the population size *P*_*i*_ of each borough, while the offset for the number of deaths given the number of cases is computed based on the number of cases observed in borough *i*. More specifically, 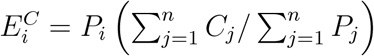 and 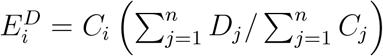. To model the number of deaths conditionally on the number of cases, the Poisson distribution is used as an approximation to a Binomial distribution with size *C*_*i*_ and probability *p*_*i*_. This approximation is reasonable because of high values of *C*_*i*_’s and small *p*_*i*_’s as estimated in an exploratory data analysis (Wakefield, 2013, section 7.3.2). In the next step, we model the log relative risks as follows:

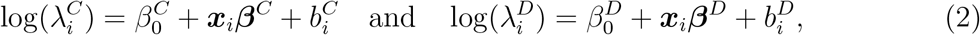

where 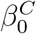 and 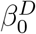 denote, respectively, the overall mean log risks of cases and deaths, ***x***_*i*_ is a vector of *K* = 3 covariates used to model both the cases and deaths associated with the *K*-dimensional vectors of coefficients, ***β***^*C*^ and ***β***^*D*^, and 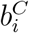 and 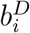 are latent random effects that accommodate whatever is left after accounting for the available covariates. Further, the inclusion of the latent random effects in both log risks’ decompositions allows for the accommodation of overdispersion which is observed in the exploratory data analysis.

We explore bivariate models for the latent random effects, 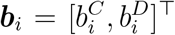. The models that we consider are special cases of the general formulation ***b***_*i*_ = Γ_*u*_***u***_*i*_ + Γ_*v*_***v***_*i*_, where the vector of independent random effects, 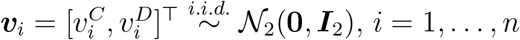, is independent of 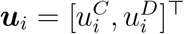, the vector of spatially structured effects following independent CAR models (Besag, 1974), and where *I*_*d*_ is the *d*-dimensional identity matrix and

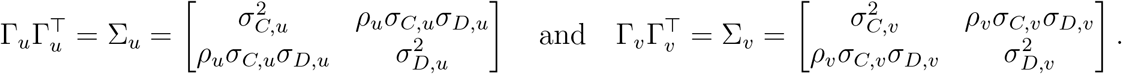

Let the CAR distributed effects 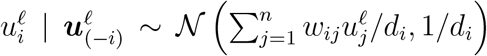, *ℓ* = *C, D*, where 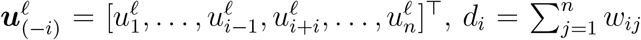 and ***W*** = [*w*_*ij*_] is the matrix of weights that defines the neighbourhood structure. Commonly, we define *w*_*ij*_ = 1 if areas *i* and *j* share a border, denoted by *i* ∼ *j*, and *w*_*ij*_ = 0, otherwise. This neighbourhood structure is used throughout this paper. Let ***D*** = *diag*(*d*_*i*_) and ***Q*** = ***D*** − ***W***, we may write ***u***^*ℓ*^∼ ***𝒩***_*n*_(**0, *Q***^−^), which is not properly defined since ***Q*** is not positive definite (Banerjee et al., 2014).

The first special case of the model for ***b***_*i*_ that we consider is one with independent random effects across the areas, denoted the IID model: 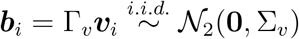. Hence, *ρ*_*v*_ denotes the correlation between the latent effects for cases and deaths. When *ρ*_*v*_ = 0, the IID joint model results in independent random effects for the cases and deaths inside each area. Further, if one assumes prior independence between the fixed effects in both models, then it does not make a difference to perform the inference about the two processes independently or jointly. In other words, there is no borrow of strength between the number of cases and deaths within boroughs. If *ρ*_*v*_ ≠ 0, this bivariate IID model allows for dependence between the deaths and cases within a particular borough.

The IID model, however, does not allow for spatial autocorrelation. Yet, one may expect the cases from neighbouring areas to be more correlated than cases from further apart boroughs, and similarly for the numbers of deaths. To adjust for this possible spatial autocorrelation, the second special case that we consider for ***b***_*i*_ is a multivariate CAR model (Lawson, 2020): ***b***_*i*_ = Γ_*u*_***u***_***i***_. One may write the joint distribution for the long vector of latent effects, 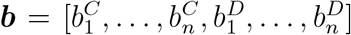, as ***b*** ∼ 𝒩_2*n*_ (**0**, Σ_*b*_ ⊗ ***Q***^−^) (Jin et al., 2007). From this joint formulation, we see that the multivariate CAR model allows for a dependence between the cases and deaths of a particular borough as well as between the counts of neighbouring areas.

The multivariate CAR model assumes *a priori* that the latent effects are necessarily distributed according to a spatial structure. To relax this assumption, we consider the multivariate extension of the BYM model (Besag et al., 1991) that allows for both unstructured and spatially structured sources of variability. This corresponds to the general formulation, ***b***_*i*_ = Γ_*u*_***u***_*i*_ + Γ_*v*_***v***_*i*_. The long vector of latent effects is distributed as ***b*** ∼ 𝒩_2*n*_(**0**, Σ_*b*_), for

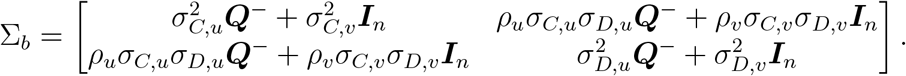

Let 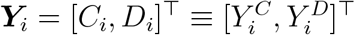. Although the marginal distribution of ***Y***_*i*_ with respect to the latent effects is not available in closed form, it is possible to obtain the marginal moments using the properties of conditional expectations and the law of total covariance. Table 1 shows the *marginal* moments of the cases and deaths obtained from the models discussed above. The detailed computation of these marginal moments is available in Appendix A. From Table 1 it is clear that the IID model is only able to capture correlation between cases and deaths within a borough. The covariance will be negative if *ρ*_*v*_ *<* 0. On the other hand, the CAR and BYM models are able to accommodate correlations both within and among neighbouring boroughs. In the CAR model, if *ρ*_*u*_ *<* 0 the correlation within and among neighbouring boroughs will be negative, and positive if *ρ*_*u*_ *>* 0. In the BYM model, there are two parameters capturing the correlation within a borough, and a negative correlation results if *ρ*_*u*_*σ*_*C,u*_*σ*_*D,u*_[***Q***^−^]_*ii*_ + *ρ*_*v*_*σ*_*C,v*_*σ*_*D,v*_ *<* 0. On the other hand, for neighbouring boroughs the correlation is captured only by the parameter *ρ*_*u*_ so that if *ρ*_*u*_ *<* 0 a negative correlation results in this case.

**Table 1:**
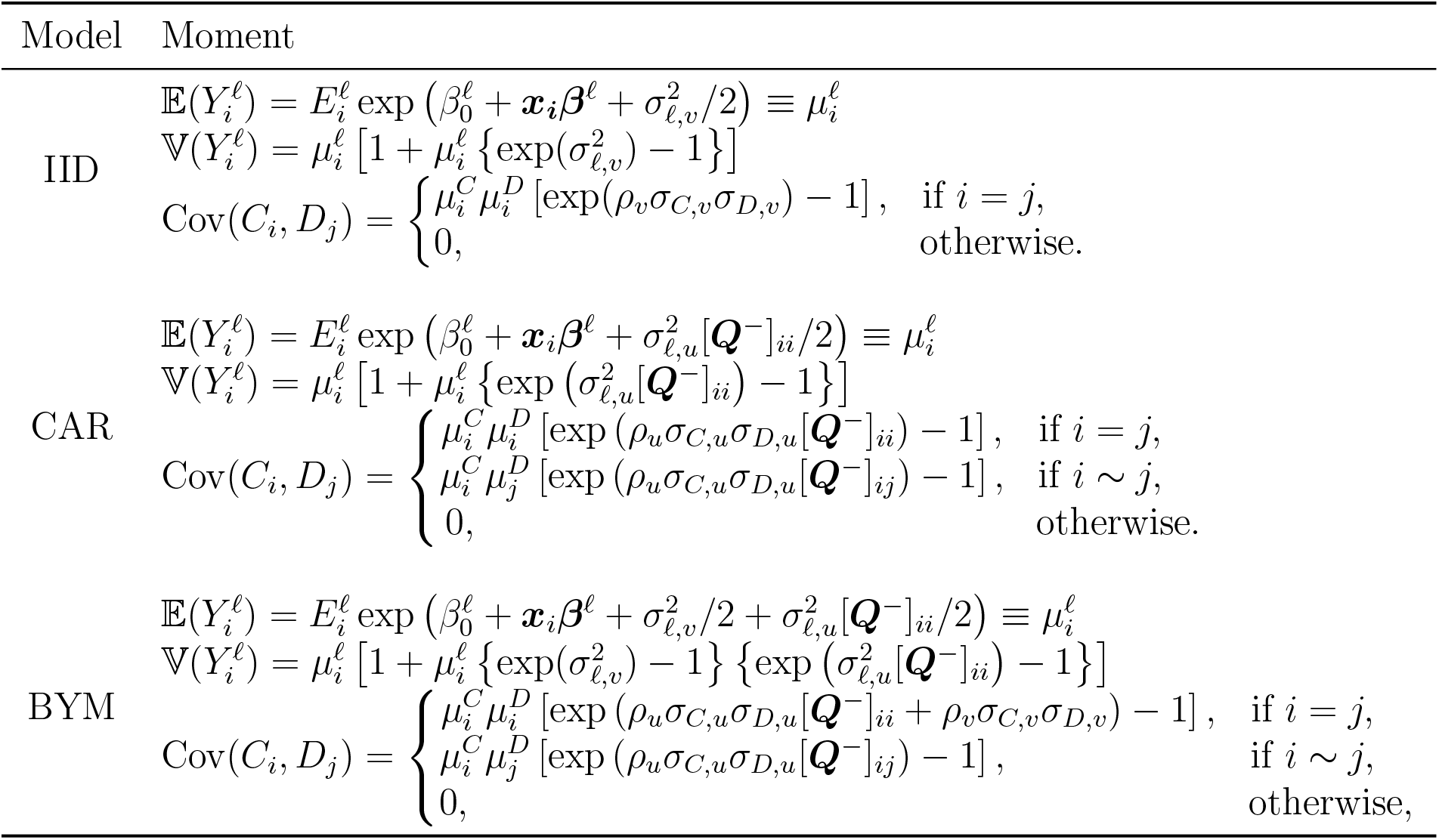
Marginal moments of 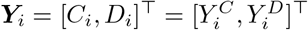, *i* = 1, …, *n*, for each model, where *ℓ* = *C, D*.

### 2.1. Investigating spatial confounding: a Bayesian alternative to Spatial+

When modelling disease risks through a Poisson model, Clayton et al. (1993) noted that covariates effects may change in the presence of spatially structured latent effects. This issue is termed spatial confounding (Reich et al., 2006). Spatial confounding corresponds to the situation where spatially structured latent effects are correlated with the covariates, resulting in biased estimates of the fixed effects (Reich et al., 2006; Page et al., 2017). Reich et al. (2006) proposed a restricted spatial regression (RSR) model to overcome this issue, by removing the collinearity between the covariates and the spatial effects. However, Khan and Calder (2020) point out that RSR models are challenging to use. Additionally to leading to a loss of computational efficiency, these models cannot be naturally extended to other spatial models than the conditional autoregressive ones. Khan and Calder (2020) also note that their use may entail an increased type-S error for both correctly and incorrectly specified models, where a type-S error occurs when a posterior confidence interval for a particular coefficient does not include 0 even though the true value is 0. Recently, Dupont et al. (2021) proposed an approach to accommodate spatial confounding called Spatial+. The proposal is to “regress away” the spatial structure from the covariates and only include the residuals from such regression in the modelling of the variable of interest. Dupont et al. (2021) suggest to use Spatial+ as a tool to investigate for the presence of spatial confounding. If after fitting a model with and without spatial effects the fixed effects estimates are similar, this might be an indication that there is no spatial confounding. The Spatial+ approach of Dupont et al. (2021) is based on generalised additive models (GAM). As pointed out by Dupont et al. (2021), because of the relationship between splines and Gaussian Markov random fields (GMRF), Spatial+ can also be used when one captures the spatial structure through a GMRF. Next, we follow Schmidt (2021) and describe how to adapt the Spatial+ approach when the latent effect follows a CAR prior distribution.

Let ***x***_*·k*_ = [*x*_1*k*_, …, *x*_*nk*_]^*T*^ be the vector for the *k*th covariate. For each covariate *k* = 1, …, *K*, we model the covariates through Gaussian distributions with latent spatial effects that follow a CAR distribution *a priori*. More specifically, let

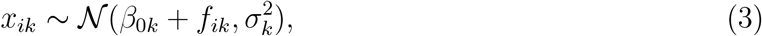

where, *a priori*, 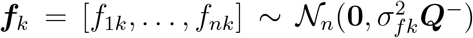. After assigning prior distributions to the parameters for the model of each ***x***_*·k*_ we follow the Bayesian paradigm and obtain a sample from the resultant posterior distribution. Then, we define *r*_*ik*_ as the posterior mean of the residual 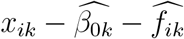, *i* = 1, …, *N, j* = 1, …, *K*, where 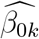 and 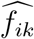 are, respectively, the point estimates (posterior means) of *β*_0*k*_ and *f*_*ik*_. Finally, each vector of potentially spatially confounded covariate, ***x***_*·k*_ is replaced by ***r***_*·k*_ = [*r*_1*k*_, …, *r*_*nk*_]^*T*^, *k* = 1, …, *K* in the models for the cases and deaths that include spatial effects.

### 2.2. Inference procedure

In the fully Bayesian framework that we consider, regardless of the prior specification of the parameters, none of the Poisson models fitted to the data result in closed form posterior distributions. Hence, we approximate these posterior distributions using computational methods, specifically, Markov chain Monte Carlo (MCMC) methods. The MCMC procedures are run in `R` using the `Nimble` package (de Valpine et al., 2017, 2021). The advantage of `Nimble` is that censored outcome values are naturally accounted for, as is necessary for the areas Baie-d’Urfé and Montréal-Ouest.

The CAR prior is not a proper distribution and is invariant to the addition of a constant (Rue and Held, 2005). This implies there is an identifiability issue between the spatially structured effects and the global mean, the intercept. To ensure the posterior distribution is proper, we impose a sum-to-zero constraint for each of the CAR distributed effects. For each of the MCMC iterations, we impose 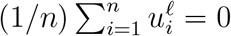. Lawson (2020) discusses how to implement the multivariate CAR model in `Nimble`. The `Nimble` codes used for this analysis are all available in Appendix C.

## 3. Results

For each of the 33 boroughs and related cities of Montreal, we have available the case and death counts due to COVID-19 that were recorded until July 25th, 2021. Let *C*_*i*_ be the case count in area *i* = 1, …, 33, and *D*_*i*_, the death count. The spatial distributions of the cases and deaths are shown in Figure 1 in Section 1. To model the cases and deaths, we consider as covariates the number of residential and long-term care centre beds, the percentage of the population with a university diploma, and the median age in each area. Figure 2 shows the maps of the variables as included in the models. The median age is scaled and the log of the number of beds is computed. Note that the log scale is used to assume a normal approximation of the number of beds. This is necessary, when fitting the Spatial+ models, as we assume that the covariates are normally distributed when adjusting for potential spatial confounding following Dupont et al. (2021). The resulting covariates from the spatial confounding adjustment are shown in Figure B.6 in Appendix B.

**Figure 2:**
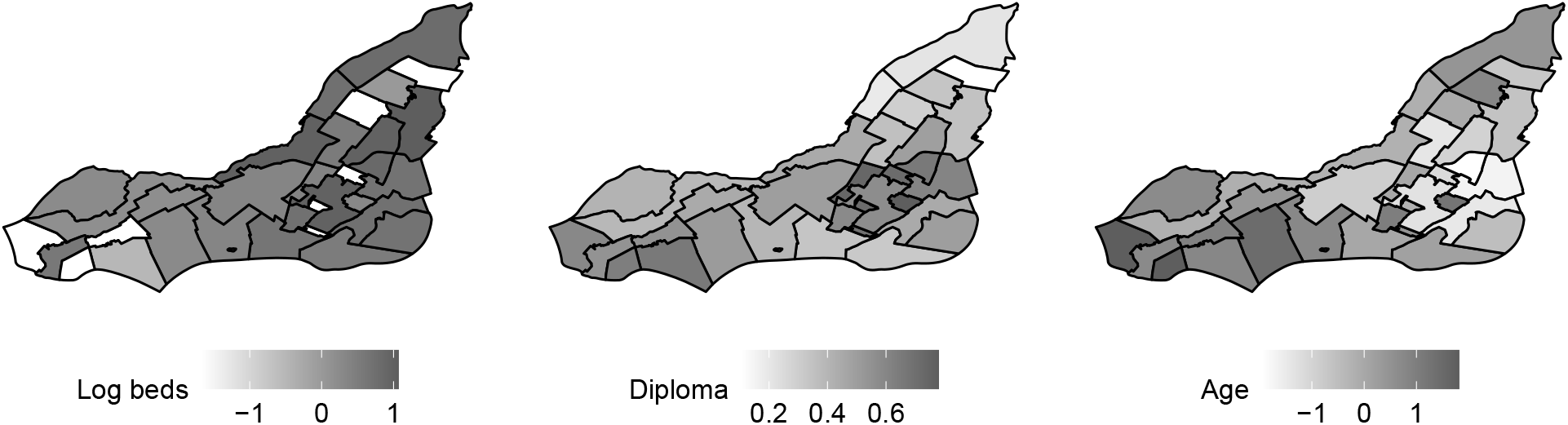
Maps of the three covariates included in the models

We are interested in assessing the association of the selected covariates with the number of cases and deaths. We are also interested in measuring the correlation between the cases and deaths inside a particular area or between neighbouring areas. To that end, we allow for a spatial autocorrelation between the cases and deaths, as described in Section 2. Hence, we fit six different Poisson models, as described in Section 2. Table 2 presents the models characteristics, namely the form of the latent effects, 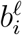, *i* = 1, …, 33, *ℓ* = *C, D*, and the covariates included. For all these models, the same priors are defined for the parameters involved in the fixed effects: 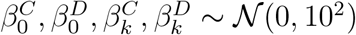, *k* = 1, 2, 3. Additionally, the relevant standard deviations that appear in the latent effects’ prior distributions are given a half-Cauchy prior, *σ*_*ℓ,v*_, *σ*_*ℓ,u*_ ∼ ℋ𝒞(0, 1), *ℓ* = *C, D*. Finally, when appropriate, the latent effects’ correlation parameters are assumed to follow a noninformative uniform prior distribution: *ρ*_*v*_, *ρ*_*u*_ ∼ 𝒰 (−1, 1).

**Table 2:** List of the fitted models to the number of cases and deaths due to COVID-19 across the boroughs of Montreal. The symbol ✓denotes which components were included in the respective model.

|  | Latent effects |  | Covariates included in the fixed effects |  |
| --- | --- | --- | --- | --- |
| | Unstructured<br>$\Gamma_v \mathbf{v}_i$ | Structured<br>$\Gamma_u \mathbf{u}_i$ | Original<br>$\mathbf{x}_{\cdot k}$ | Adjusted for Spatial Confounding<br>$\mathbf{r}_{\cdot k}$ |
| Simple | - | - | ✓ | - |
| IID | ✓ | - | ✓ | - |
| CAR | - | ✓ | ✓ | - |
| BYM | ✓ | ✓ | ✓ | - |
| CAR+ | - | ✓ | - | ✓ |
| BYM+ | ✓ | ✓ | - | ✓ |

The models are fitted through the `R` package `Nimble` (de Valpine et al., 2021). The MCMC procedure consists of 2 chains of 800,000 iterations each, with a burn-in period of 400,000 iterations and a thinning factor of 160. An elliptical slice sampler is used for the regression coefficients, as these are normally distributed *a priori*. This sampler was chosen to efficiently sample from these posterior full conditionals. Metropolis-Hastings algorithms are used for the independent random effects in the IID, BYM and BYM+ models. For the spatial effects included in the CAR, BYM, CAR+, and BYM+ models, the default CAR normal sampler from Nimble is used. The chains have mixed well, as assessed by the trace plots, effective sample sizes and the statistic proposed by Gelman and Rubin (1992). The reason for the need of these long chains seems to be related to the possible collinearity between the variable diploma and the models that include a spatial effect. The IID model and the ones that accommodate spatial confounding showed convergence before reaching the burn in of 400,000 iterations. For consistency, all models were fitted with the same number of iterations.

The posterior summaries for the fixed effects’ coefficients are presented in Figure 3 for each model. The solid circles correspond to the estimated posterior means, while the segments are the limits of the 95% posterior credible intervals. The first row corresponds to the fixed effects in the log relative risk of cases and the second row, to the ones in the log relative risk of recorded deaths due to COVID-19. Clearly, the inclusion of random effects in each of the equations change the importance of each of the covariates. The covariate beds is negatively associated with the log relative risk of cases under the Simple model and as a random effect is included in the model, zero falls within the 95% posterior credible interval, suggesting that the number of beds is not associated with the cases. For the log relative risk of deaths, on the other hand, the association with the number of beds is strictly positive. This is expected as the majority of deaths in Quebec are connected to residential and long-term care centres.

**Figure 3:**
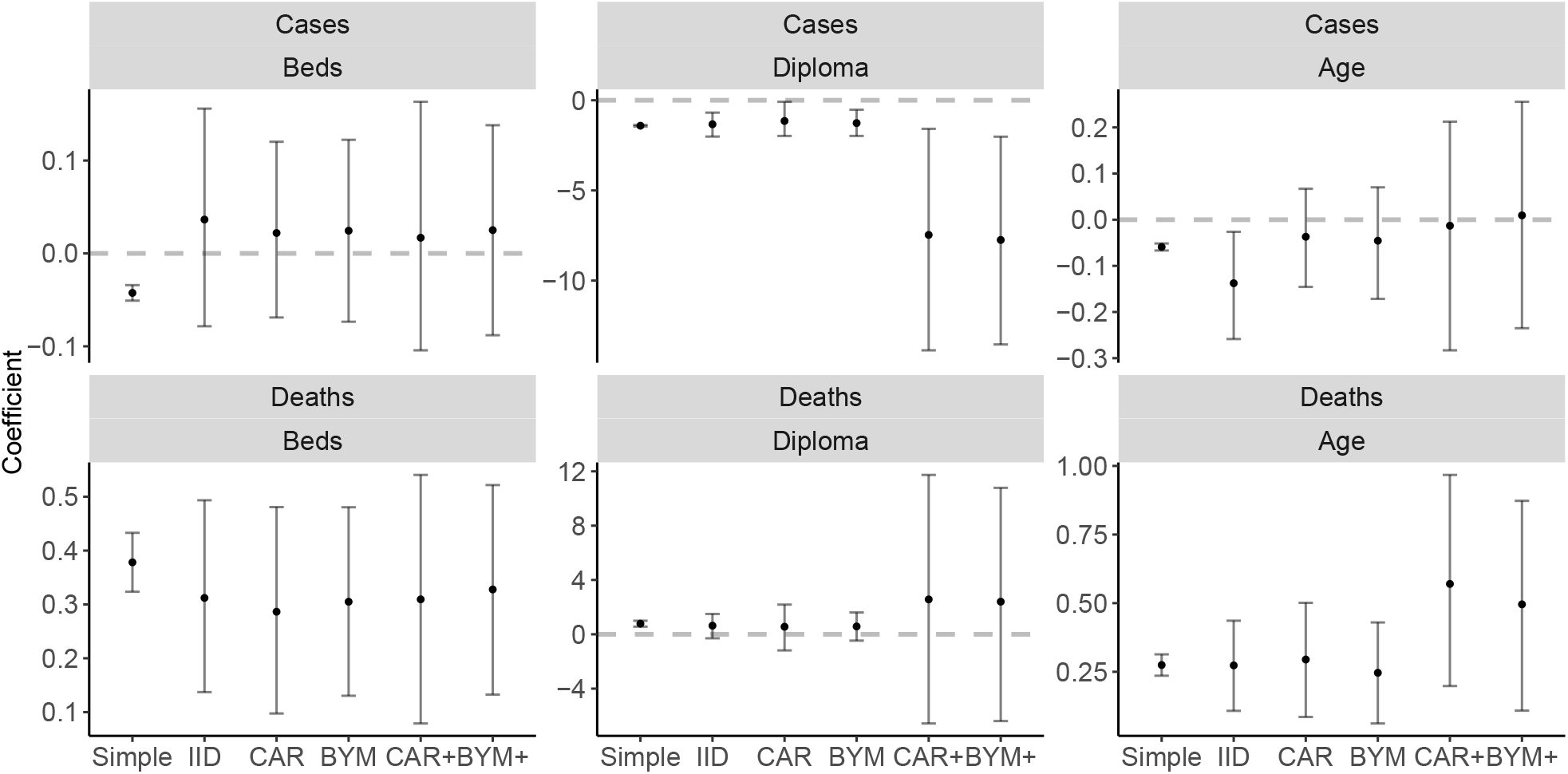
Posterior summaries for the model coefficients across all the fitted models. Solid circles: posterior means; Vertical lines: 95% posterior credible intervals; Dashed line: indicates no association.

The variable diploma is negatively associated with the log relative risk of cases. The range of the 95% posterior credible interval is much wider under the CAR+ and BYM+ models than the ones obtained under the IID and CAR models. Although diploma results in a positive association with the log relative risk of death under the Simple model, once a random effect is included, zero is within the 95% posterior credible of the coefficient associated with diploma. This suggests that the average educational level of the borough is not associated with the risk of death.

For the median age of the borough, the Simple and IID models suggest a negative association with the log relative risk of cases and a positive association with the log relative risk of deaths. For the log relative risk of cases once a spatial structure is included in the model, zero falls within the 95% posterior credible interval. This is also true for the CAR+ and BYM+ models which account for spatial confounding.

We model the cases and deaths jointly in order to borrow strength from the different recordings both inside each area (e.g., IID model) and between areas (e.g., CAR model). Figure 4 shows the posterior summaries (posterior mean and posterior 95% credible interval) for the correlation coefficients, *ρ*_*v*_ and *ρ*_*u*_, respectively included in the latent effects’ IID, CAR, CAR+, BYM or BYM+ models. The BYM and BYM+ yield posterior 95% credible intervals that include 0 for *ρ*_*u*_ and *ρ*_*v*_. On the other hand, the IID, CAR and CAR+ result in negative correlations between the latent effects for the cases and for the deaths ([−0.73,−0.08], [−0.72,−0.07] and [−0.76,−0.02], respectively). This suggests that modelling the cases and deaths jointly is adequate. Note that for the BYM and BYM+ models, if the posterior 95% credible limits for *ρ*_*u*_ and *ρ*_*v*_ include zero, this does not necessarily imply that the marginal correlations will also include zero. From Table 1, it is clear that under models BYM and BYM+ the correlation between cases and deaths within and across boroughs also depends on the latent effects’ standard deviations as well as the spatial structure. Figure B.7 in Appendix B shows the posterior summaries for the intra-borough correlation between cases and deaths for each model. Although *ρ*_*u*_ and *ρ*_*v*_ have posterior credible intervals that include zero, the BYM model yields strictly negative intervals in 19 boroughs.

**Figure 4:**
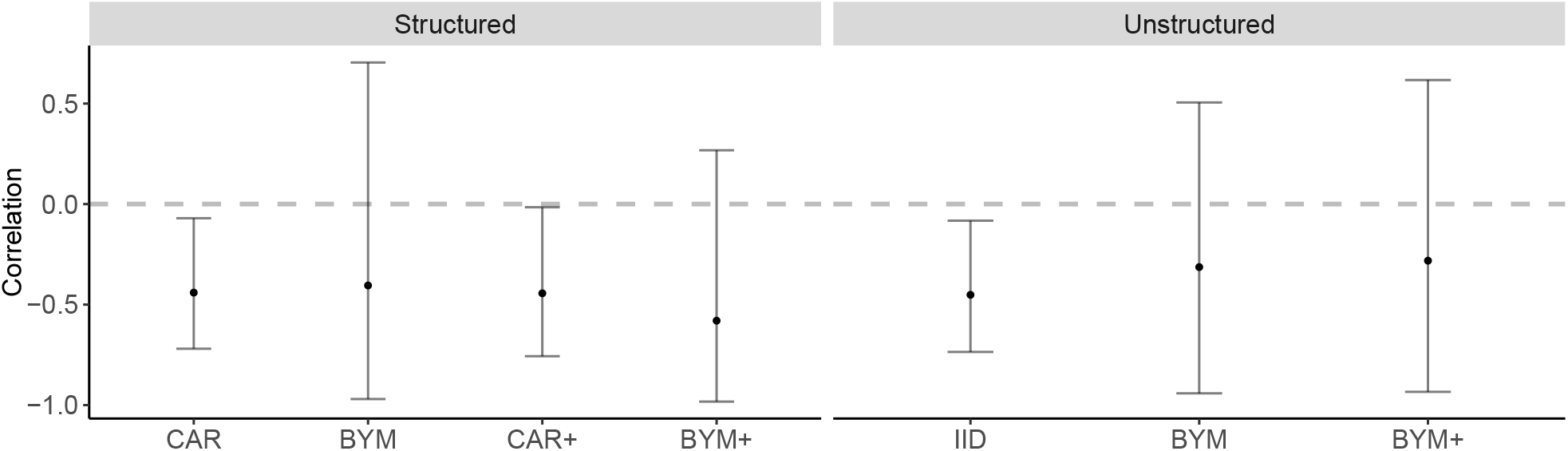
Posterior summaries for the correlation coefficient of the latent effects. Solid circles: posterior means; Vertical lines: 95% posterior credible intervals. Dashed line: *ρ*_*u*_ = 0 and *ρ*_*v*_ = 0.

Another goal of this analysis, other than to investigate the association of the log relative risks of cases and deaths with the available covariates, is to estimate the correlation between the cases and deaths within and across boroughs. The posterior summaries for the intra-borough correlations between cases and deaths obtained for each model are available in Appendix B, in Figure B.7 and the posterior means for the correlation between areas estimated under the CAR, BYM, CAR+, and BYM+ models are available in Appendix B, in Figure B.8, Figure B.9, Figure B.10 and Figure B.11, respectively. We note that all the estimated correlations are negative. Figure 5 maps the posterior means of the correlations between cases and deaths inside each borough for each model that include latent effects. The IID model does not allow for a correlation across boroughs but does accommodate an intra-borough correlation (see Table 1). All the models estimate negative correlations between the cases and deaths on average *a posteriori*. For example, the correlation’s posterior mean in Senneville is −0.2 for the CAR model and −0.1 for the BYM model. In Côte-des-Neiges-Notre-Dame-de-Grâce, the posterior mean is −0.4 for the CAR+ model and −0.3 for the BYM. For the IID, CAR and CAR+ models, the correlations’ posterior 95% credible intervals (see Figure B.7 in Appendix B) are always negative. In Figure 5, there does not seem to be a spatial structure in the negative posterior means of the intra-borough correlations between the cases and deaths due to COVID-19. Additionally, Figure B.12 in Appendix B maps the relative risks of cases and deaths as estimated by each model. The negative correlations between cases and deaths can be visualized in this Figure B.12. For example, the southwest region of Montreal has low estimated risks of cases but high estimated risks of deaths for each model.

**Figure 5:**
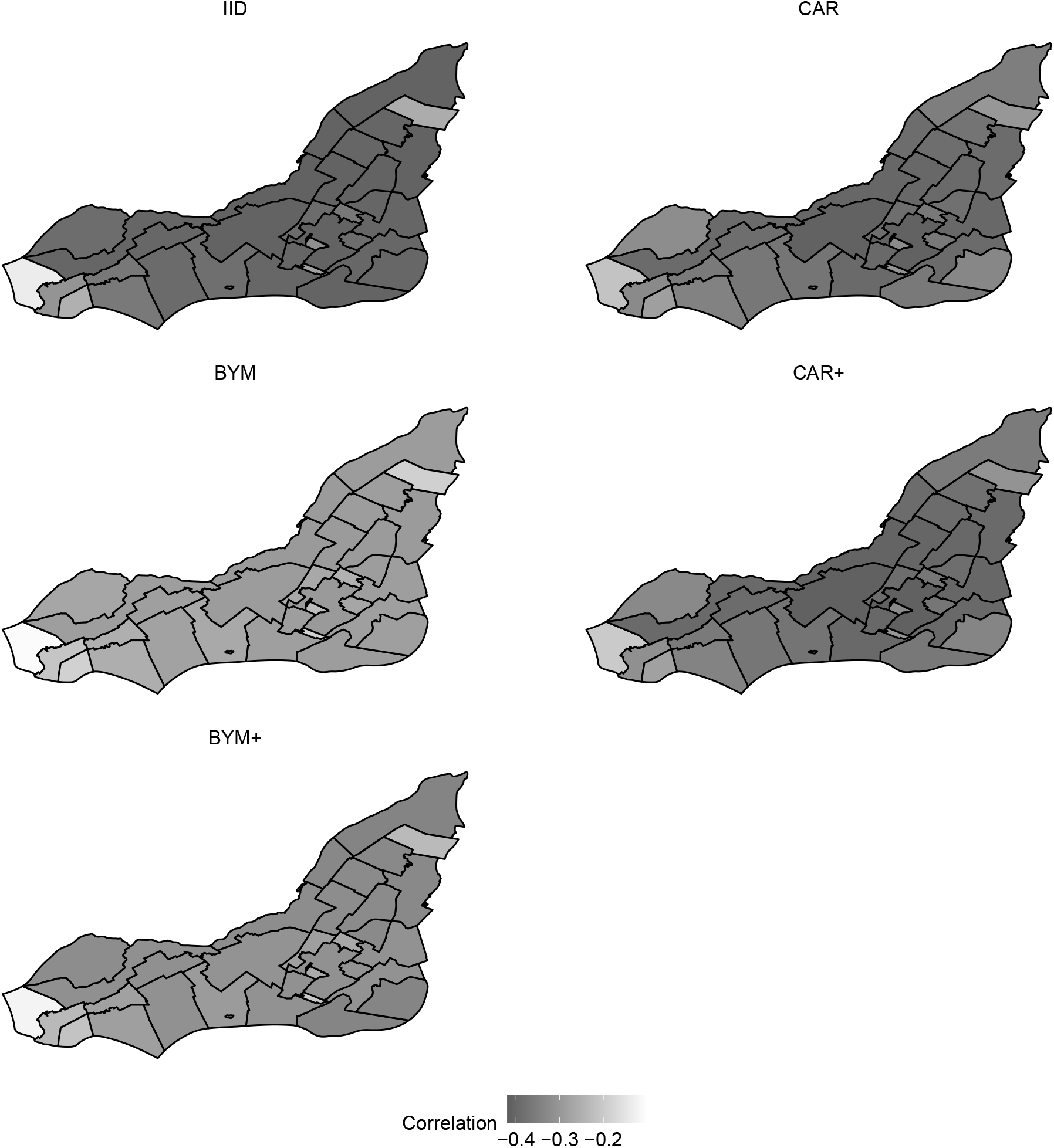
Maps of the posterior means of the intra-borough correlation between cases and deaths for each model. These correlations were estimated based on the equations for the covariance under each model shown in Table 1.

Finally, we compare the models’ performances using the WAIC (Watanabe and Opper, 2010) in Table 3. In terms of WAIC, for which smaller values are preferred, the Simple model is the least adequate among the fitted ones, with a value of 3,320 and 265 effective number of parameters. This suggests that there is a need for latent effects in the modelling of cases and deaths due to COVID-19 in Montreal. However, there does not seem to be a need for a spatial structure in these random effects as the CAR, BYM, CAR+, and BYM+ all perform worse than the IID model. The IID model yields a WAIC of 205 whereas the others yield a WAIC of 212, 225, 243 and 227, respectively.

**Table 3:** Values of WAIC and effective number of parameters for each fitted model. The smallest value indicates the best model among fitted ones (in italics).

|  | Simple | IID | CAR | BYM | CAR+ | BYM+ |
| --- | --- | --- | --- | --- | --- | --- |
| WAIC | 3320.1 | *205.0* | 212.2 | 224.9 | 242.7 | 227.1 |
| $p_W$ | 265.2 | *57.2* | 60.7 | 67.2 | 75.3 | 68.0 |

## 4. Discussion

In this paper, we analyse the case and death counts due to COVID-19 across the 33 boroughs of Montreal, as of July 25th, 2021. We fit six different models with conditional Poisson distributions for the cases and for the deaths in each borough. The areal means are decomposed in the log scale as the sum of an offset, fixed effects, and latent random effects. One model is fitted without latent effects. We then allow for a correlation between the cases’ latent effects and the deaths’ latent effects within an area. In one model, we assume that the latent effects are independent across the areas, whereas in the other four models, we accommodate a potential spatial autocorrelation between the latent effects of neighbouring areas. In two of these models, we set multivariate CAR and BYM priors for the latent effects. In the final two models, we first adjust the covariates for potential spatial confounding following Dupont et al. (2021) and then consider again multivariate CAR and BYM priors for the latent effects.

Interestingly, the posterior summaries for the fixed effects parameters are reversed between the cases and deaths components of the models. Regarding the log number of beds, the coefficient has posterior 95% credible intervals that include zero for the cases (through the IID, CAR, BYM, CAR+, and BYM+ models) but the posterior credible intervals are strictly positive for the deaths. We used the number of beds in CHSLDs as a proxy for the healthcare infrastructure in Montreal. This result suggests that this covariate has no link with the recorded number of cases but that, given the number of cases, an increased number of beds corresponds to a higher risk of death due to COVID-19. This result is consistent with the COVID-19 situation in Montreal and in the province of Quebec. In particular, residential and long-term care centres were the epicentre of the COVID-19 crisis in Quebec (Hsu et al., 2020; Institut national de santé publique du Québec, 2021). The associations between the percentage of the population with a university diploma and the cases and deaths due to COVID-19 in Montreal are also different for each outcome considered. There does not seem to be a relationship between the percentage of people with diploma and the risk of death due to COVID-19, given the cases, for the IID, CAR, BYM, CAR+, and BYM+ models. On the other hand, each model results in negative posterior credible intervals for the effects of the diploma on the number of cases. The diploma variable is used as a proxy to the socio-economic situation of each borough in Montreal. This result seems to be aligned with other studies that also found that level of education is negatively associated with the log risk of cases (see e.g., Hawkins et al. (2020)). Finally, the risk of COVID-19 cases does not seem to be associated with age for the CAR, BYM, CAR+, and BYM+ models whereas older populations seem correlated with higher risks of death due to COVID-19. Once again, this result agrees with the current knowledge on COVID-19: all may contract COVID-19, but younger people are less likely to die from this disease (Williamson et al., 2020).

As the importance of the covariates changed when a latent spatially structured random effect was considered, we also investigated if there is spatial confounding by fitting a Bayesian version of the method proposed by Dupont et al. (2021). We find that the log number of beds does not seem to be spatially confounded whereas the diploma and median age do. As suggested by Dupont et al. (2021), we believe there is an interest in adjusting for spatial confounding even when there is no intuition of spatial confounding, as their method can be used as a test for the presence or absence of such confounding. However, in terms of WAIC, the model with only IID latent effects seems to perform the best among the fitted ones. This suggests that there is no need to account for spatially structured latent effects. Note that the adjustment for spatial confounding is unnecessary in this IID model.

We believe that there is an interest to the data analysis conducted in this paper, as we are able to provide an estimate of the correlations between the cases and deaths due to COVID-19 within, and across, boroughs. Fitting separate models for cases and deaths, or even joint models that assume parameters to be independent *a priori*, such as in the Simple model, impose independence between cases and deaths. Our proposed approach allows the data to drive the inference procedure, and if there is correlation between the two outcomes, this can be estimated through the marginal correlations computed from the results shown in Table 1. Note that in terms of WAIC, we find that the Simple model performs worse than the five other models that include latent effects which allow for marginal correlations between cases and deaths both within and across boroughs. Finally, we find that the correlation between the cases and deaths due to COVID-19 within and between boroughs is always negative. This may be due to the aggregation of cases and deaths over time. We only have available aggregated data, which may yield different correlations than what would be observed at a certain time or through a temporal analysis. Looking at aggregated provincial time series of cases and deaths due to COVID-19, the number of cases and deaths were positively correlated at the beginning of the pandemic. We hypothesise that the estimated negative correlation between cases and deaths are related to different policies that were instituted throughout the pandemic and, moreover, due to the fact that people started being vaccinated. For instance, in the province of Quebec, the number of cases increased from 573 to 1683 between March 15th, 2021, and April 15th, 2021, whereas the vaccination coverage for the population aged 80 and older increased from 57.9% to 91.1% in the same period (Institut national de santé publique du Québec, 2021). It can be observed in the aggregated time series that, in this period, the daily number of deaths somewhat stabilised at low numbers.

In this paper, we used the median age of the boroughs as a proxy to the age population profile. If data for different age groups are available, one could consider models for case and death counts for each age stratum, allowing for different effects of the available variables and correlation parameters across the different age groups. In this framework, one would need to carefully consider the inclusion of the number of long-term care home beds in the various strata.

## Data Availability

The cumulative numbers of cases and deaths due to COVID-19 recorded across the boroughs of Montreal that are analysed in this study are made publicly available by the "Direction regionale de sante publique". The population sizes, the median age and the percent of the population aged 25-64 with a university diploma come from the 2016 Canadian Census, which is publicly available. The number of beds in long-term care homes across the boroughs are obtained from a public database by the "Ministere de la Sante et des Services Sociaux". The shape files used to map the data are also publicly available.

https://santemontreal.qc.ca/population/coronavirus-covid-19/situation-du-coronavirus-covid-19-a-montreal/

http://ville.montreal.qc.ca/portal/page?_pageid=6897,68149755&_dad=portal&_schema=PORTAL

https://publications.msss.gouv.qc.ca/msss/document-001644/

https://santemontreal.qc.ca/population/coronavirus-covid-19/situation-du-coronavirus-covid-19-a-montreal/

## Appendix A Detailed computation of the covariance between cases and deaths

In this section, we develop the general expression for the correlation between the cases and deaths inside each area and between areas. Recall the general formulation of the model described in section 2 for *C*_*i*_, the number of cases in area *i, i* = 1, …, *n*, and *D*_*i*_, the death count:

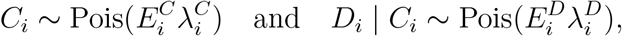

where Pois stands for the Poisson distribution and where 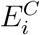 and 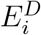 are offsets. We further decompose the relative risks, 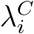 and 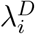, in the log scale as follows:

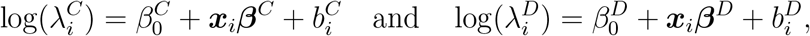

for 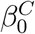 and 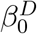 the mean log risks and for ***x***_*i*_ the vector of *K* covariates associated to the coefficients ***β***^*C*^ and ***β***^*D*^. The latent random effects 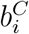 and 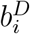 may be defined in the general form:

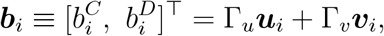

where 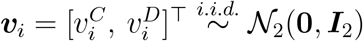 is independent of 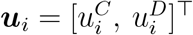, for ***u***^*ℓ*^∼ 𝒩 (**0, *Q***^−^), *ℓ*= *C, D*, with ***I***_*d*_, the *d × d* identity matrix, ***Q***, the matrix defining the spatial structure, and with

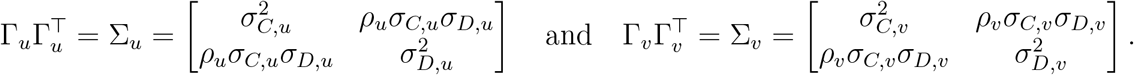

Hence, the distribution for the long vector 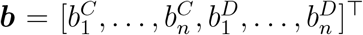 is ***b*** ∼ 𝒩_2*n*_(**0**, Σ_*b*_), for

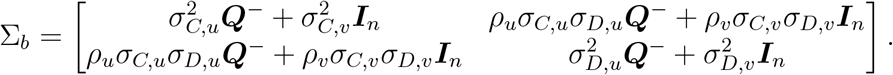

Given the fixed effects, we can compute the *marginal* covariances between cases and deaths, marginalising over the random effects:

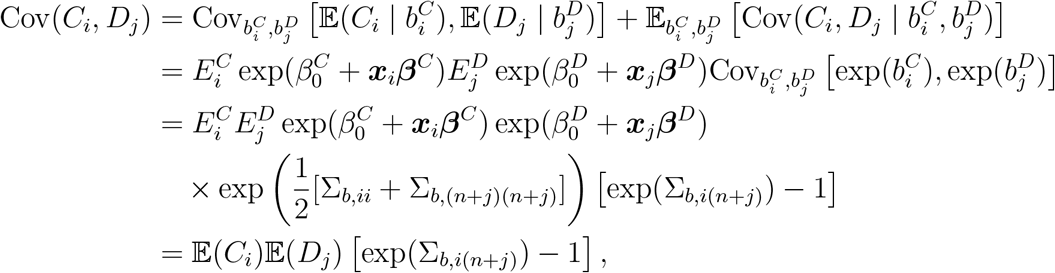

with marginal expectation for the cases 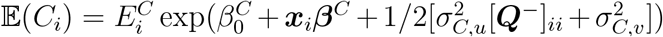 and for the deaths 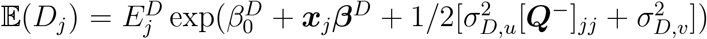. Hence, inside a particular area *i*, for *i* = *j*, we have

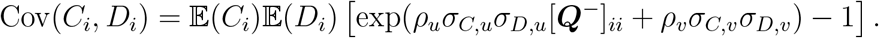

Across areas, if *i* and *j* are not neighbouring areas, then Cov(*C*_*i*_, *D*_*j*_) = 0 and if *i* ∼ *j*, we obtain

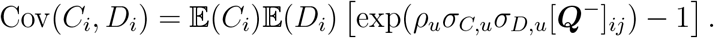

## Appendix B Additional results from the analysis of cases and deaths due to COVID-19 in Montreal

The maps of the covariates after adjusting for the spatial confounding are shown in Figure B.6. Figure B.7 shows the posterior summaries for the intra-borough correlation between the cases and deaths, as estimated by the IID, CAR, BYM, CAR+, and BYM+ models. Figures B.8, B.9, B.10 and B.11 plot the posterior means for the correlations between boroughs. The posterior means for the relative risks from each model are mapped in Figure B.12.

**Figure B.6:**
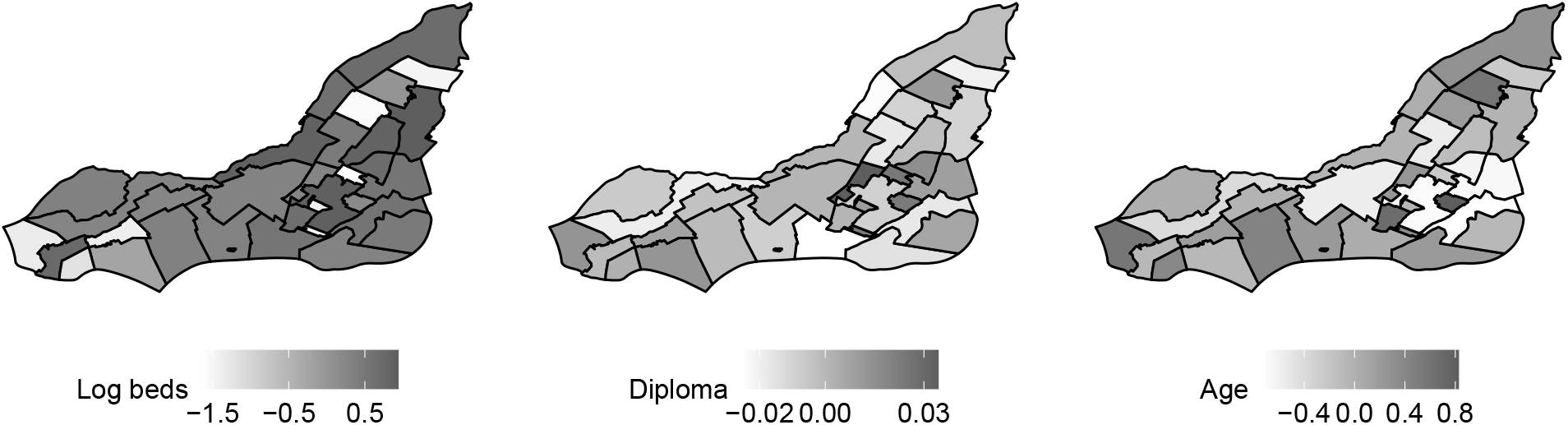
Maps of the three covariates included in the models after adjusting for spatial confounding

**Figure B.7:**
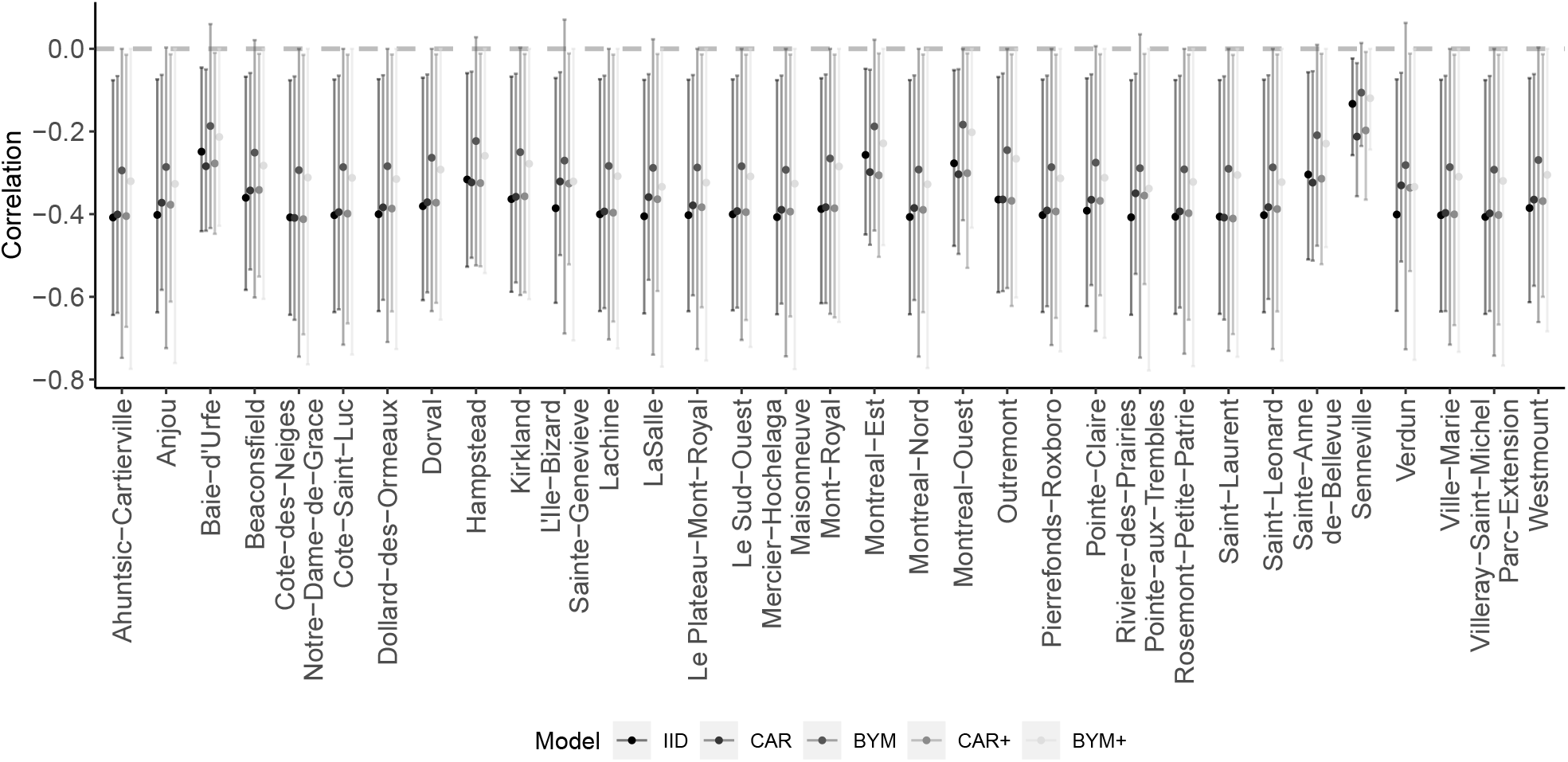
Posterior summaries for the intra-borough correlation between cases and deaths for each of the IID, CAR, BYM, CAR+, and BYM+ models

**Figure B.8:**
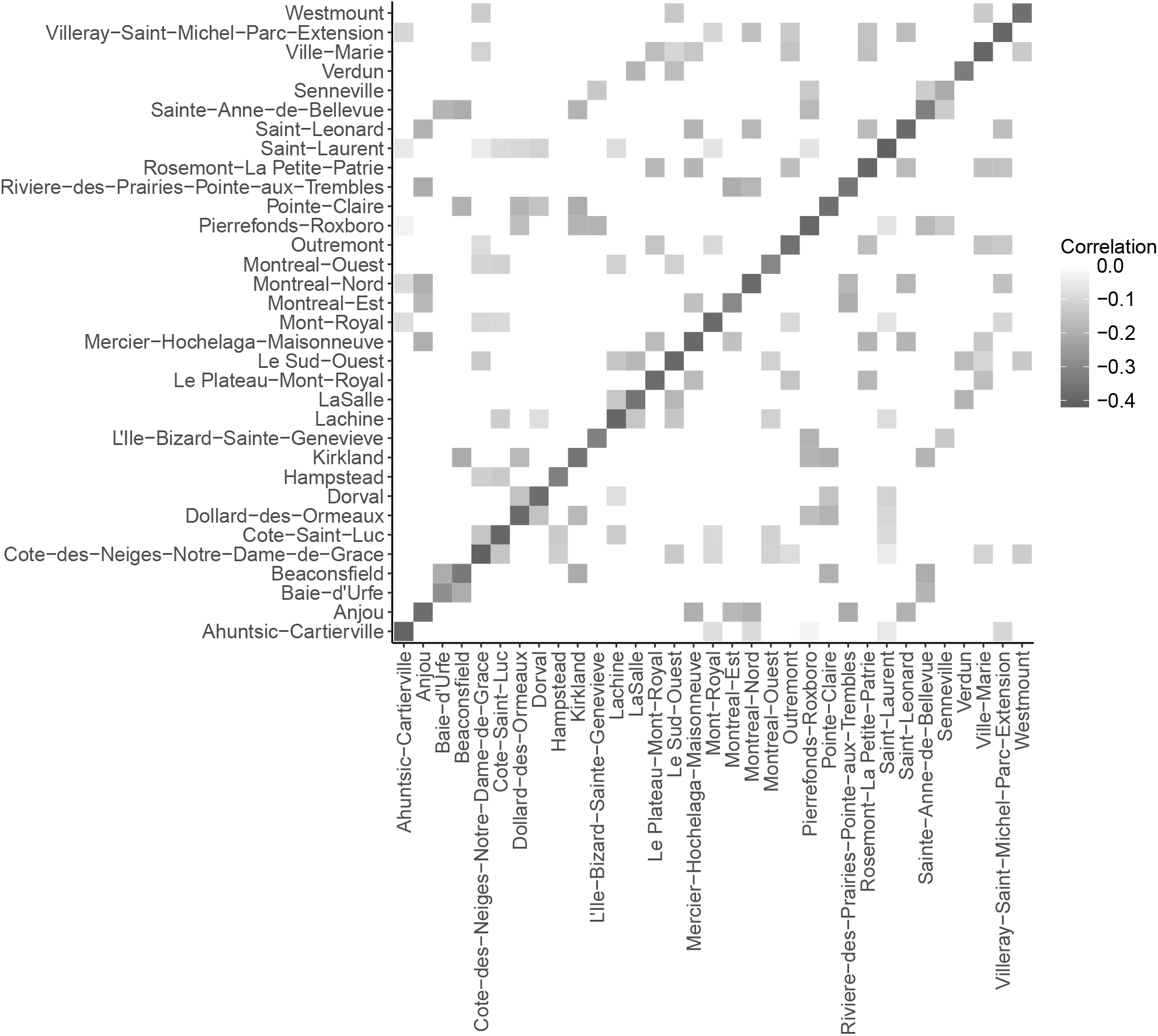
Posterior means of the correlation between deaths and cases inside and between the borough for the CAR model

**Figure B.9:**
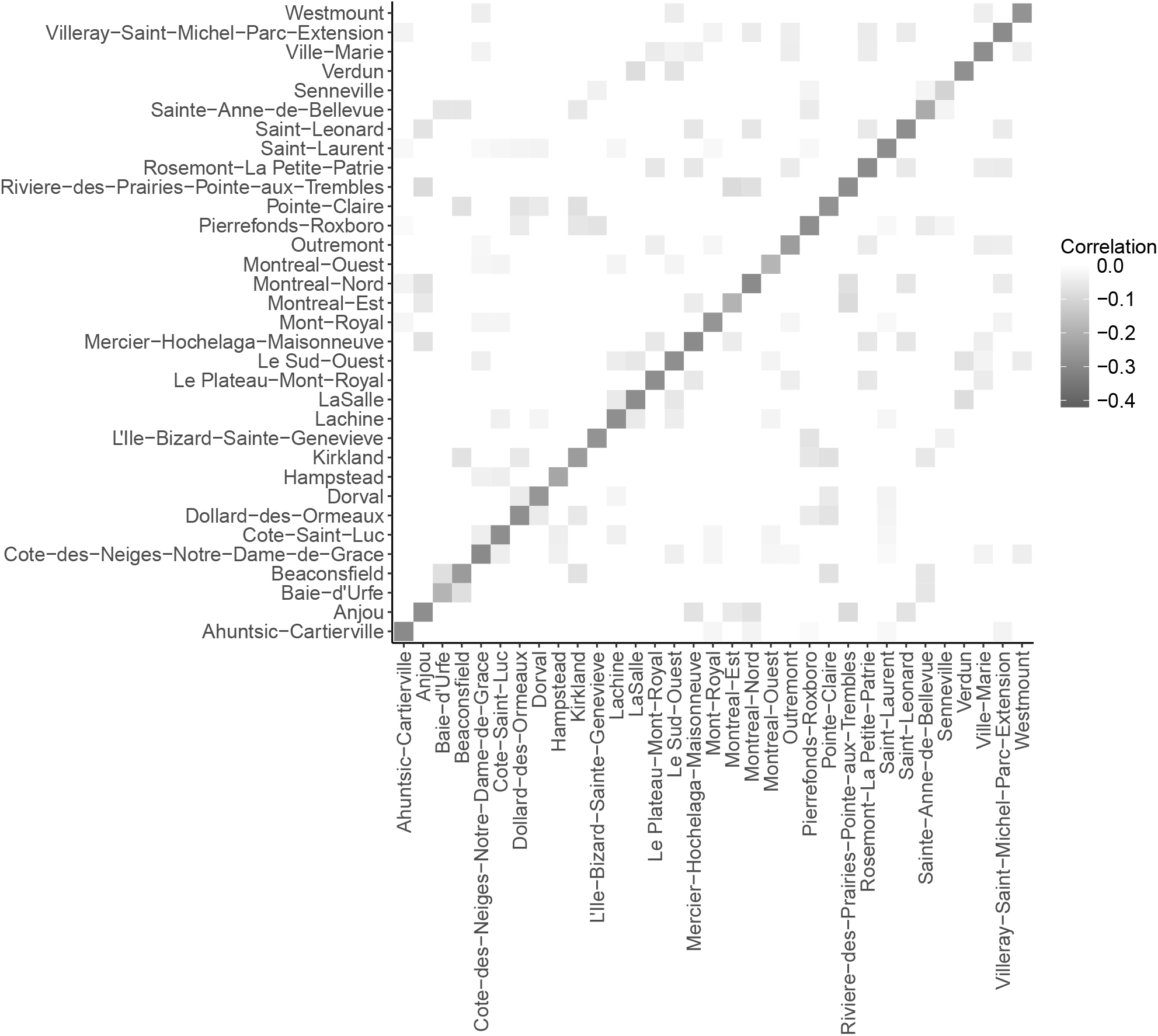
Posterior means of the correlation between deaths and cases inside and between the borough for the BYM model

**Figure B.10:**
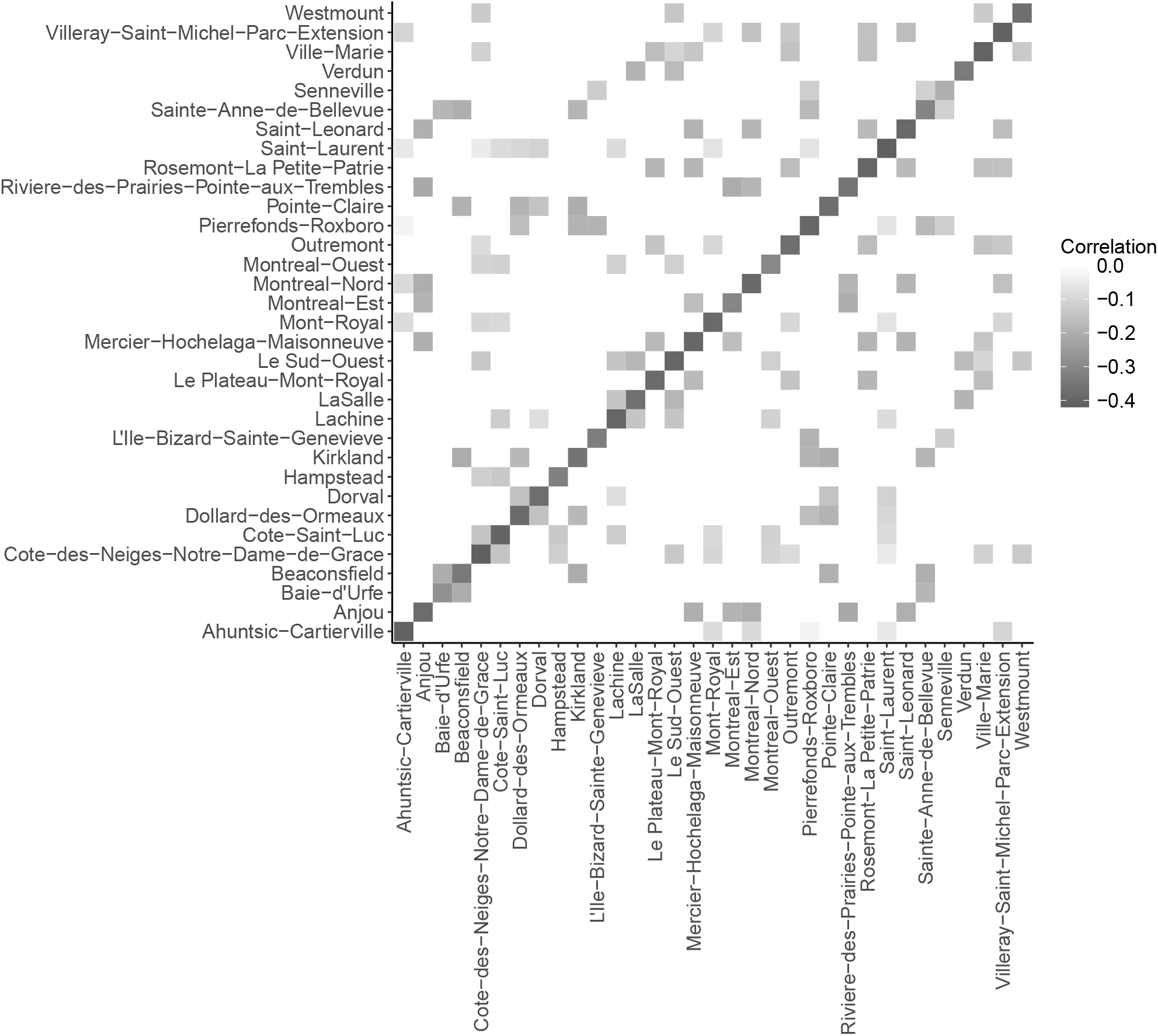
Posterior means of the correlation between deaths and cases inside and between the borough for the CAR+ model

**Figure B.11:**
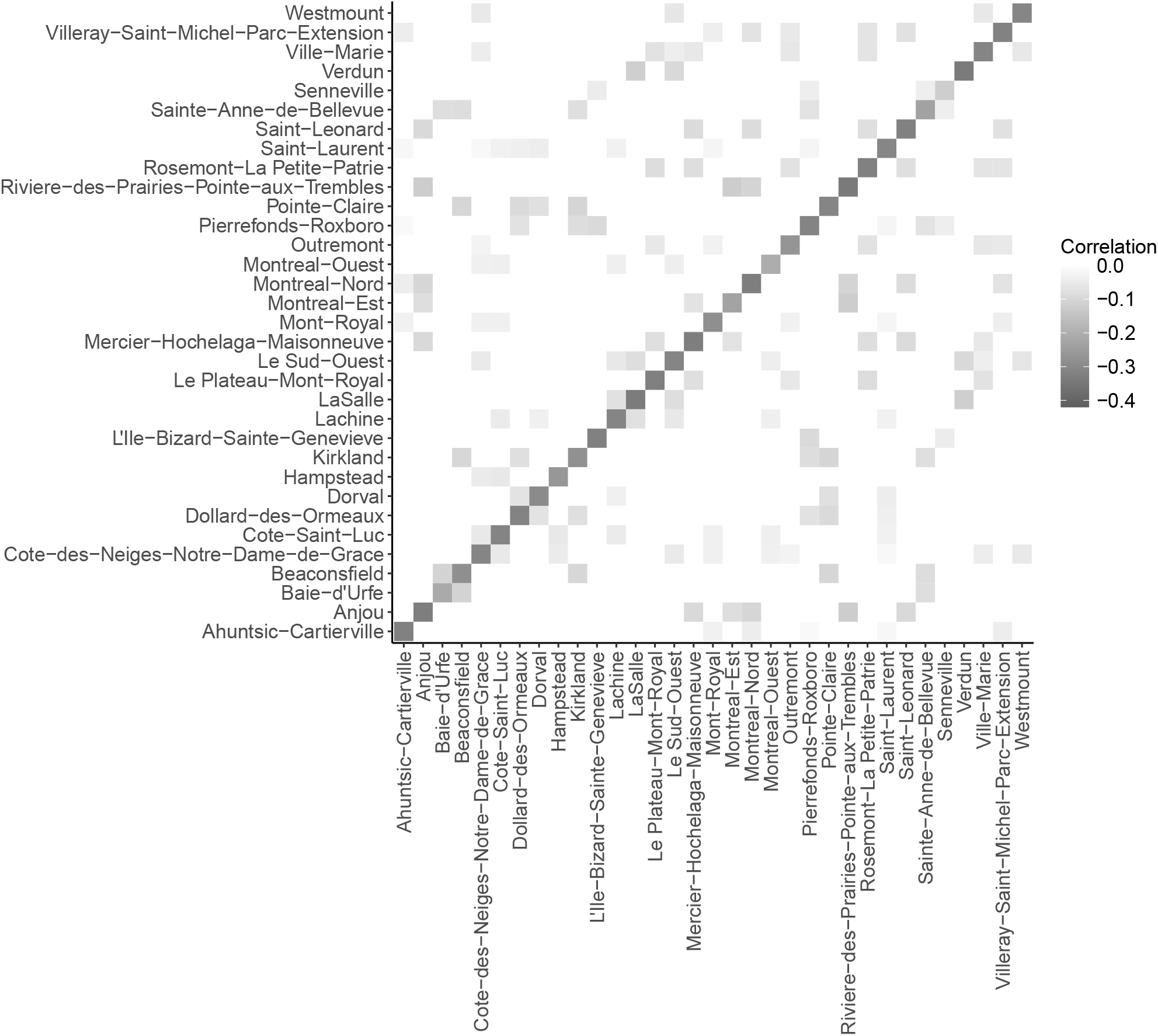
Posterior means of the correlation between deaths and cases inside and between the borough for the BYM+ model

**Figure B.12:**
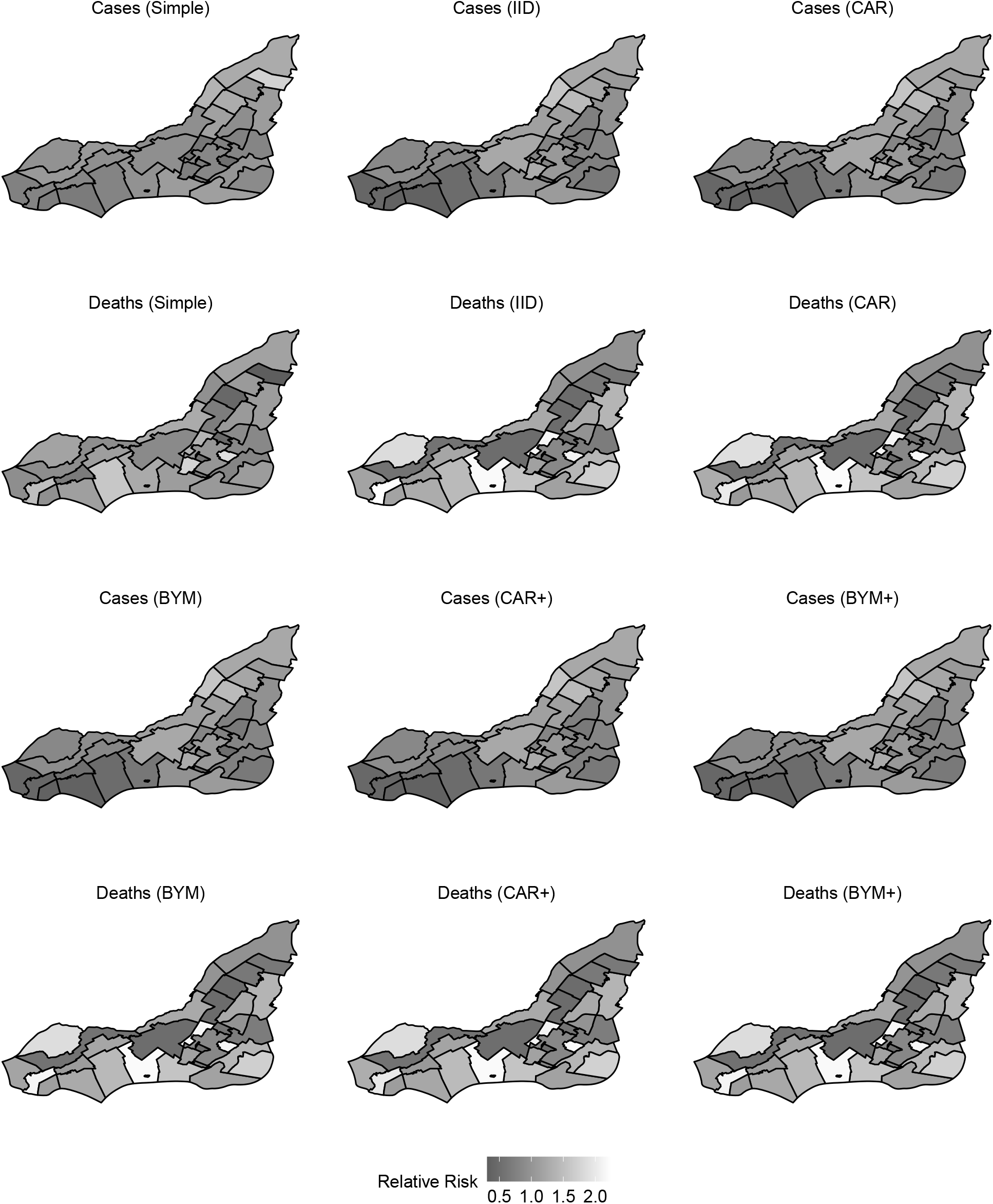
Maps of the posterior means for the relative risks estimated from the six models for the cases and deaths. Rows 1 & 2: Simple, CAR and IID models; Rows 3 & 4: BYM, CAR+, and BYM+ models.

## Appendix C Nimble codes

The Nimble codes used to perform the analyses presented in this paper are listed below.

**Listing 1:**
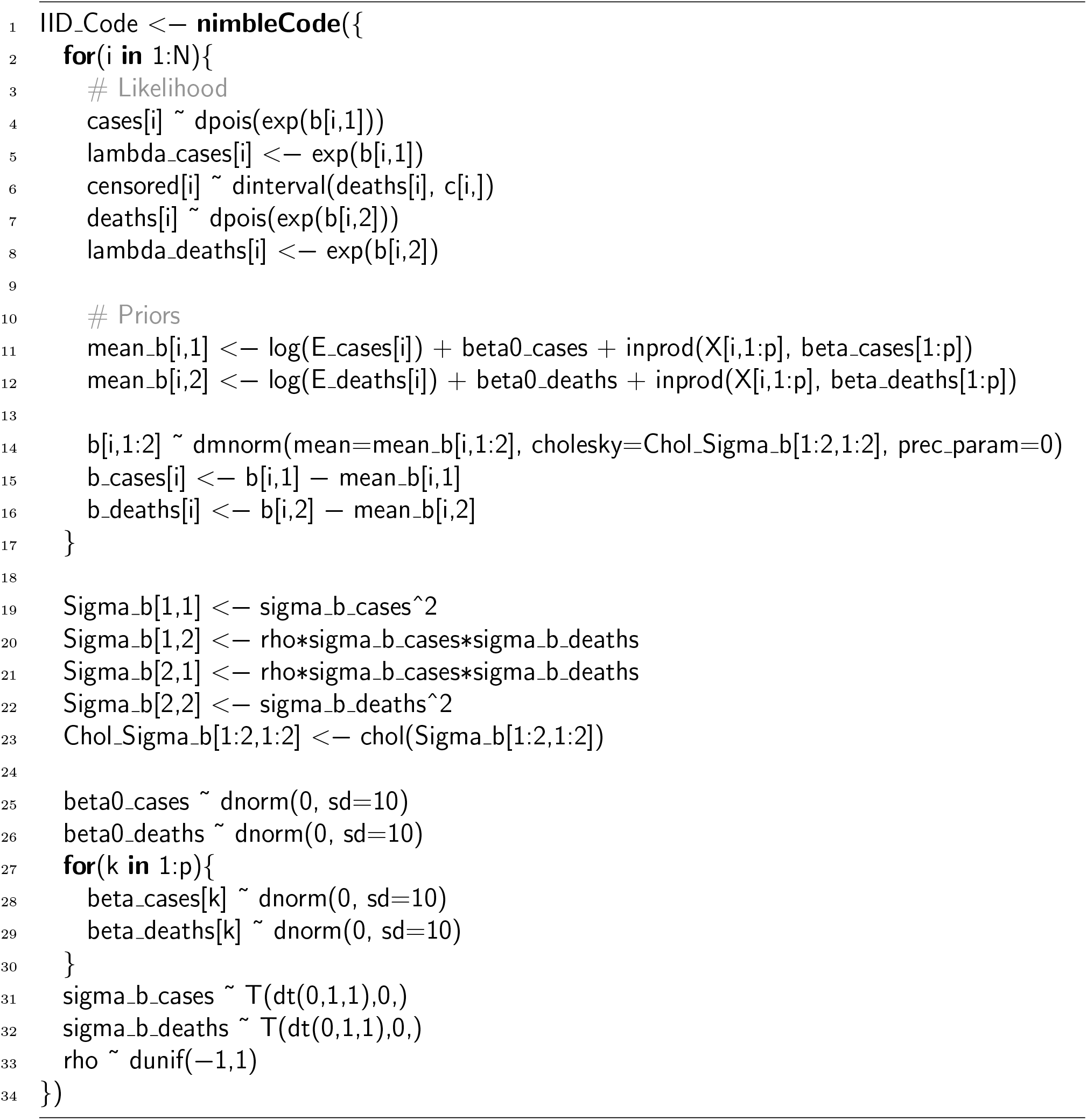
Nimble code for the joint model with independent latent effects (IID)

**Listing 2:**
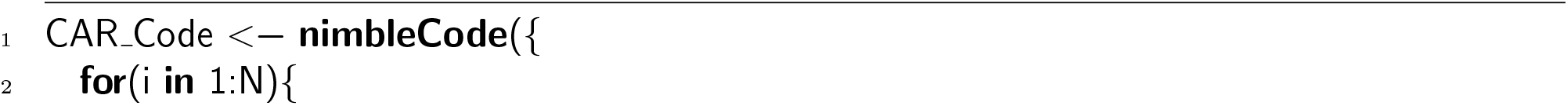
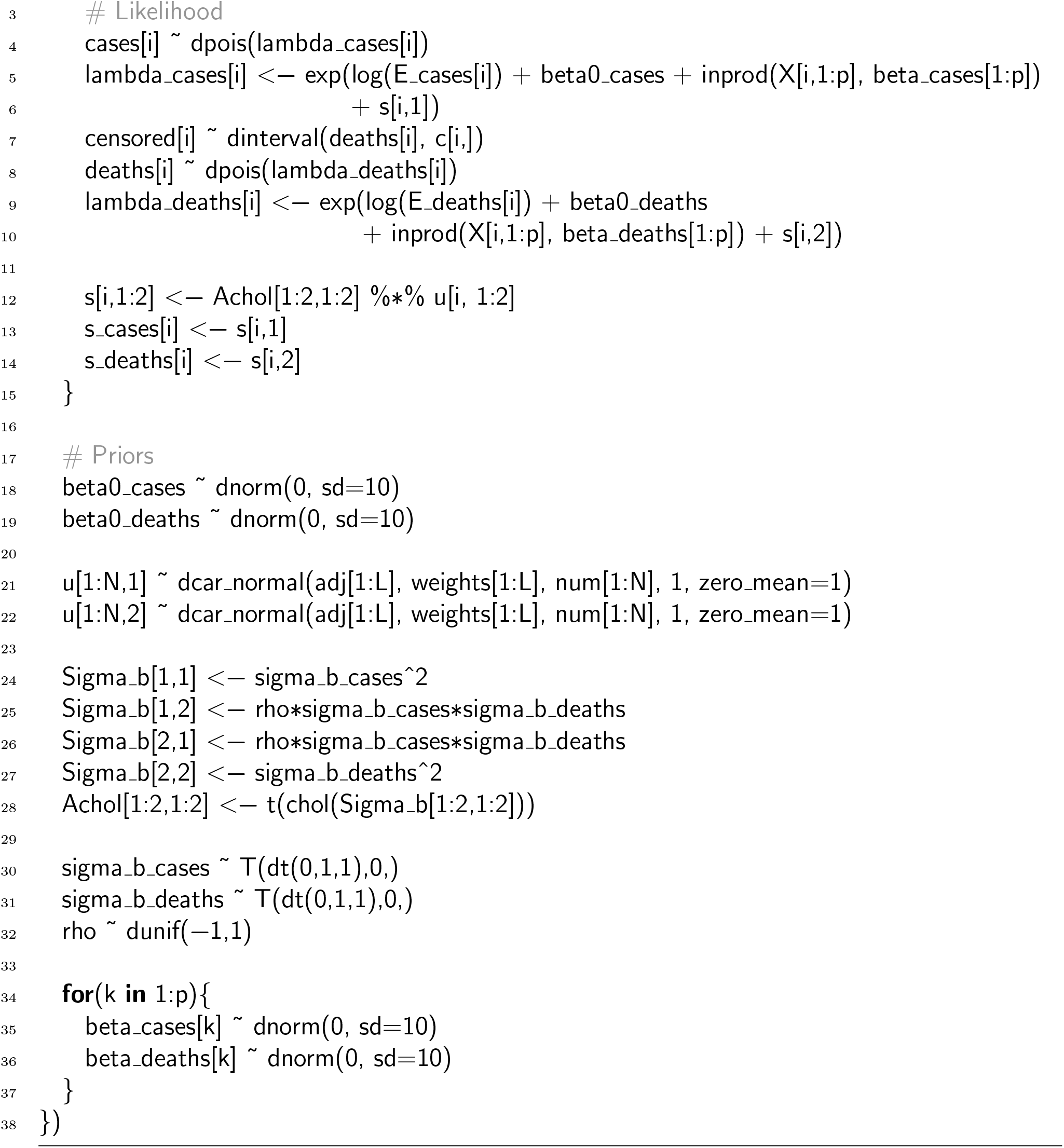
Nimble code for the joint model with spatial latent effects (CAR)

**Listing 3:**
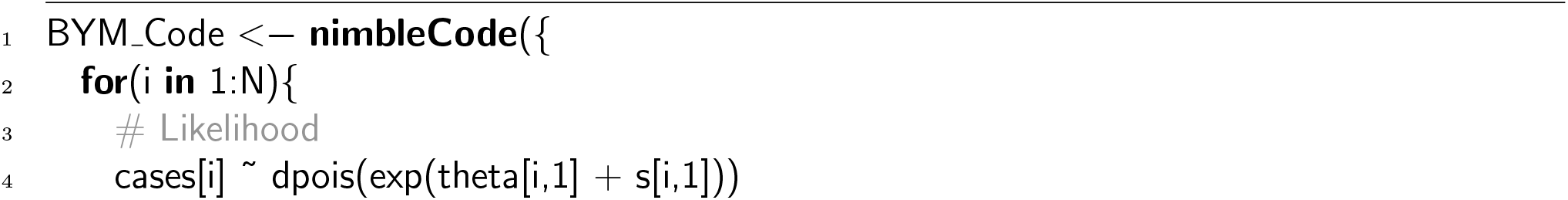
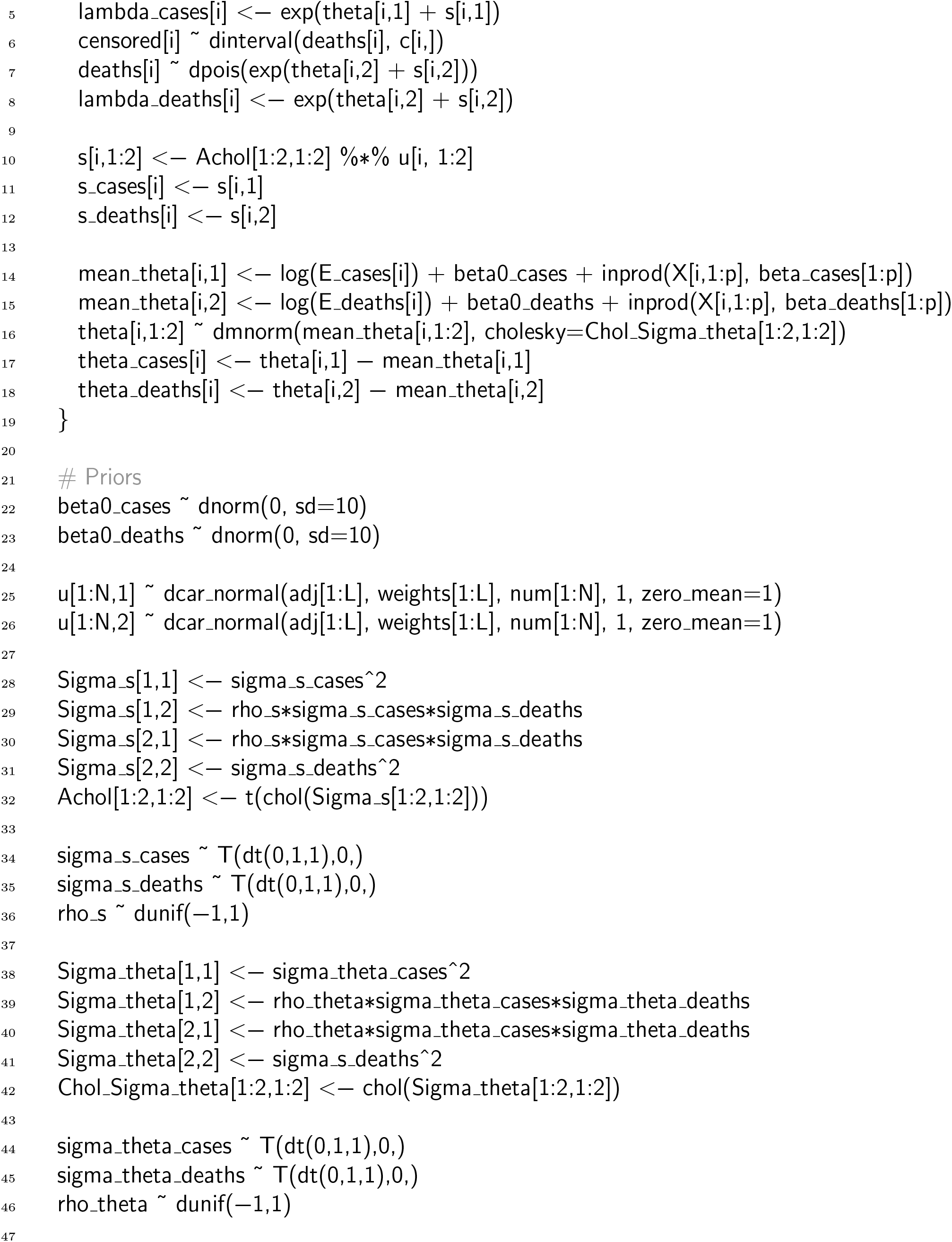
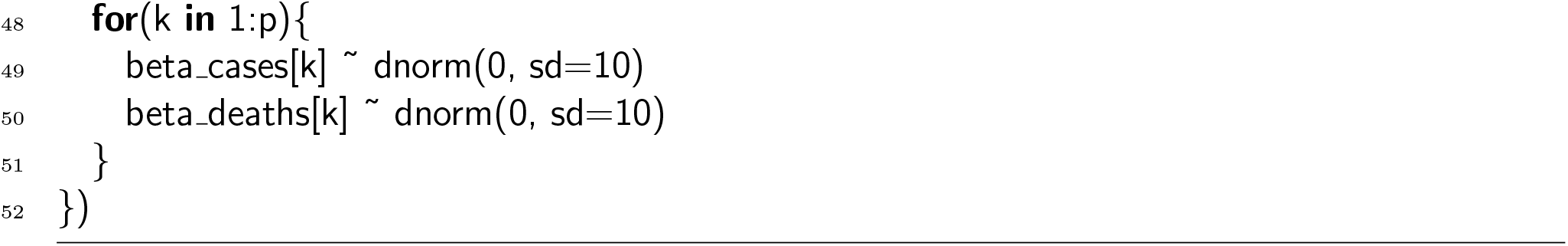
Nimble code for the joint model with spatial and independent latent effects (BYM)

